# Prevalence and predictors of adverse birth outcomes among postnatal mothers in Dodoma: Hospital-based analytical cross-sectional study

**DOI:** 10.64898/2025.12.16.25342343

**Authors:** Kina Shafii Msuya, Fabiola Vicent Moshi, Walter C Millanzi

## Abstract

**Background:** Adverse birth outcomes are a common health problem consisting of several health effects on postnatal mothers. Maternals with one or more adverse birth outcomes are at greater risk for maternal mortality. Despite ongoing government efforts to improve maternal care, the magnitude of adverse birth outcomes is a remarkable health concern. Maternal mortality is still a major public health challenge in Tanzania.

**Objective:** This study aimed to assess the prevalence and predictors of adverse maternal outcomes among postnatal women.

**Methods:** The study employed a quantitative, hospital-based cross-sectional design in the Dodoma region. Multiple sampling methods were used to choose the health facilities, and simple random sampling was used to select study participants. The study involved 422 out of 425 postnatal mothers with a 99.3 percent response rate. Data from respondents was collected using a questionnaire and a documentary review of antenatal cards and patient files. IBM SPSS version 25 for data analysis, employing both descriptive and inferential statistics.

**Results:** The findings showed that participants who were emotionally abused were more likely to develop eclampsia, and 4.275 times more likely to develop anaemia compared to participants who were not emotionally abused (AOR = 3.864; 95%CI: 1.766–8.456: *p* = 0.001), (AOR = 3.079; 95%CI: 1.514, 6.262: *p =* 0.002), and (AOR = 4.275; 95%CI: 2.020, 9.045: *p =* <0.001) respectively. Participants who had a history of stillbirth were (AOR = 3.811; 95%CI: 1.696, 8.562: *p = 0.001).* Participants who had their delivery conducted at GA <37 weeks were more likely to have anaemia GA ≥37 weeks (AOR = 2.846; 95%CI: 1.327, 6.102: *p* = 0.007). In addition, participants who had <4 ANC visits were 5.091 more likely to have an infection ≥4 ANC visits (AOR = 5.091; 95%CI: 1.093, 23.717: *p = 0.038)*

**Conclusion:** These findings underscore the importance of comprehensive ANC services that prioritize mental health support, address social determinants of health, and ensure equitable access to healthcare. Interventions to increase ANC attendance, address abuse, and improve healthcare accessibility, particularly for those living far from health facilities, are essential to improve maternal and neonatal health outcomes.

## Introduction

Adverse birth outcomes are significant public health issues that can arise during pregnancy, labor, and the postpartum period possibly affecting mothers and or their newborn babies. Some of these outcomes are postpartum hemorrhage (PPH), infections during pregnancy, anemia, eclampsia and low birth weight, preterm birth, stillbirth, and anemia respectively (1).

While substantial progress has been made in maternal and neonatal health globally, ongoing efforts are being made to ensure that all women and newborns receive care as needed. Globally, the maternal mortality rate has dropped from approximately 385 deaths per 100,000 live births in 2015 to 216 in 2017, which indicates substantial progress. Significant disparities exist in Sub-Saharan Africa, with the highest maternal mortality rate accounting for about 66% of global maternal deaths (2).

On a global scale, 2.4 million deaths of newborns occur in the first 28 days, and a quarter occur within the first three weeks. The neonatal mortality rate decreased from approximately 37 deaths per 1000 live births in 2015 to 17 in 2020. However, the progress varies significantly across the regions. Sub-Saharan Africa continues to experience the highest neonatal mortality rate, with more than 30 deaths per 1000 live births (3).

Pregnancy, childhood, and the postnatal period should bring positive experiences by ensuring mothers and their babies reach their full potential for health and well-being. Healthy women and neonates are those whose pregnancies last a full nine months with normal labor, healthy babies weighing at least 2500g–3500g with no defects, and a mother who feels well throughout the entire nine months (4).

Postnatal women with excellent health can contribute significantly to newborn breastfeeding to provide optimal nutrition to their neonates, impacting their newborn development milestones. Furthermore, it will have an impact on supporting family dynamics through positive role models, community development engagement, advocacy, and return work to increase the family’s and country’s income (5).

However, postnatal mothers encounter several health-related challenges during the postnatal period, such as breastfeeding issues, urinary incontinence, depression, puerperal psychosis, post-traumatic pain, stress disorders, sleep disorders, and adverse maternal birth outcomes (6).

Additionally, the post-delivered mother may suffer from extensive perineal tear, acute mastitis, persistent lower abdominal pains, endometriosis, and other adverse maternal outcomes such as postpartum hemorrhage, hypertensive disorders, postpartum anemia, uterine rupture, and maternal death (7).

The magnitude of adverse birth outcomes is a remarkable health concern worldwide, especially in low and middle-income countries (8). Globally, the prevalence of maternal adverse birth outcomes is 28.3% (6). Sub-Saharan Africa’s prevalence of maternal adverse outcomes accounts for 35% (9–11).

Factors contributing to maternal-adverse birth outcomes in developed countries include the maternal experience of interpersonal discrimination that may lead to a range of adverse maternal outcomes, including various physiological markers of stress. (Sonderlund et al., 2021).

Factors contributing to adverse neonatal outcomes in developing countries include alcohol use during pregnancy, consumption of Arabic tea, substance use, and cigarette smoking (Bayih et al., 2021). In Sub-Saharan Africa, factors such as premature rupture of membranes, a lack of standard fetal heartbeat monitoring, the presence of antepartum hemorrhage, and hypertension have been identified (12). In Tanzania, a study conducted to assess factors associated with low birth weight in Dar-es-Salaam revealed that malaria and HIV during pregnancy are factors that contribute to neonatal adverse outcomes (13).

Moreover, the current maternal mortality ratio in Tanzania is 104 maternal deaths per 100,000 live births (14). The major direct causes of maternal death were eclampsia 34.0%, obstetrical hemorrhage 24.6%, maternal sepsis 16.7%, anemia 14.9%, and cardiovascular disorders 14.0% (14).

SDGs 3.1 which aim to reduce maternal mortality to less than 70/100,000 by 2030, describe global efforts to improve these rates (15). Strategies included addressing disparities in the quality and availability of sexual, reproductive, maternal, and newborn healthcare; guaranteeing universal health coverage for comprehensive sexual, reproductive, maternal, and newborn healthcare; addressing all causes of maternal death, morbidities related to reproduction, and disabilities associated with it; building the health system to meet the needs and priorities of women and girls; and ensuring accountability to improve care quality and equity (16).

Tanzania has implemented several strategies to lower the rates of maternal and newborn deaths. These include collaborating closely with other partners, decentralizing and emphasizing high-quality services, setting up efficient monitoring and evaluation systems, promoting the sustainability of high-quality maternal and newborn care, educating providers, and offering payment for services (17). Based on the information provided it seems that the problem of maternal adverse outcomes exist. The available data suggests that underdeveloped nations, such as Tanzania, face a more serious issue with unfavorable maternal outcomes than wealthy nations. Tanzania’s healthcare system has long regarded maternal and newborn health as a crucial aspect. The Ministry of Health created several national policies and recommendations to address the outcomes of maternal and neonatal births (18). All of these recommendations aimed to enhance the delivery of maternity and newborn healthcare services. By reducing the rate of newborn deaths to 20 per 1,000 live births, the Tanzanian government has successfully decreased the maternal mortality rate from 556 per 100,000 live births to 104 live births. However, the apparent decline in death rates does not align with the targets of the SDGs (19).

The development of ANC guidelines, ensuring the availability of necessary medications and equipment, creating an essential newborn care training package, providing Emergency Obstetric and Newborn Care (EmONC) training, improving medical infrastructure, and improving administrative responsiveness are among the initiatives and interventions that the Ministry of Health has modified and created.

Dodoma is among the top ten regions in the country with a high number of maternal and neonatal deaths that is 104/100,000. Therefore, this study aimed to determine the prevalence and predictors associated with adverse maternal outcomes in the Dodoma region.

## Material and Methods

### Study area

The study area was conducted in three selected hospitals in the Dodoma region: Dodoma Referral Reginal Hospital, ST. Gemma Hospital, and Uhuru Hospital. According to the 2022 Tanzania Demographic and Health Survey, the region has a total of 497 health facilities (402 dispensaries, 69 health centers, and 26 hospitals).

The region’s population is estimated to be 3,085,625 according to a national census survey, males are 1,512, 760 and females 1,572,865 (URT, 2023). Dodoma is among the top ten regions in the country with higher maternal and newborn deaths (20). The population growth rate was 3.2 (10).

## Study Approach and Design

### Study Approach

This current study deployed a quantitative approach to obtain participant data through a questionnaire and documentary review of patient fails and ANC cards.

### Study design

The researcher used an analytical cross-sectional study design to determine the prevalence of adverse outcomes and the relationship between relevant adverse birth outcomes.

### Inclusion criteria

All postnatal mothers were admitted within 0 – 28 days post-delivery.

### Exclusion criteria

Postnatal mothers aged below 18 years were not included in this study and those who delivery by caesarian section.

### Sample size estimation

A Kish Leslie (1965) will be used to calculate the required sample size.

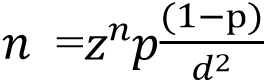

Where by:

n = Minimum sample size,

z = Confidence interval 1.96,

d = marginal error,

The proportion of 50% was used to determine a minimum sample size since no other study was conducted in the area.

n = 1.96^2^ x 0.5(1-0.5)

0.05^2^

n = 383

The sample size was 383 respondents. By adjusting for non-response will be 10%: N = 383/1-0.1 The total sample size is 425

### Sampling Technique

A mixed sampling methods was employed to get districts, hospital and participants. A simple random sampling by lottery method was used to select two districts out of 7 districts in Dodoma region. The selected districts were Dodoma city and Chamwino district.

In Dodoma city, a regional referral hospital was purposively selected because it is the referral hospital which receive patients from different parts of the region, thus making it potential for larger number of cases. Then a simple random sampling method was used to select one hospital in each district out of average of 5 for both districts. The selected hospital were St. Gema in Dodoma city and Uhuru in Chamwino district.

The proportionate sampling was then used to get number of participants from each hospital using a formula n= Ni/Nt x n

Where:

ni=Requiredd numbers required of postnatal mothers in the selected facility

Ni= Total number of postnatal mothers in the facility chosen.

Nt= Total number of postnatal women in all selected health facilities

n= Estimated sample size

The obtained number of participants in each facility is shown in table 1.

**Table 1:**
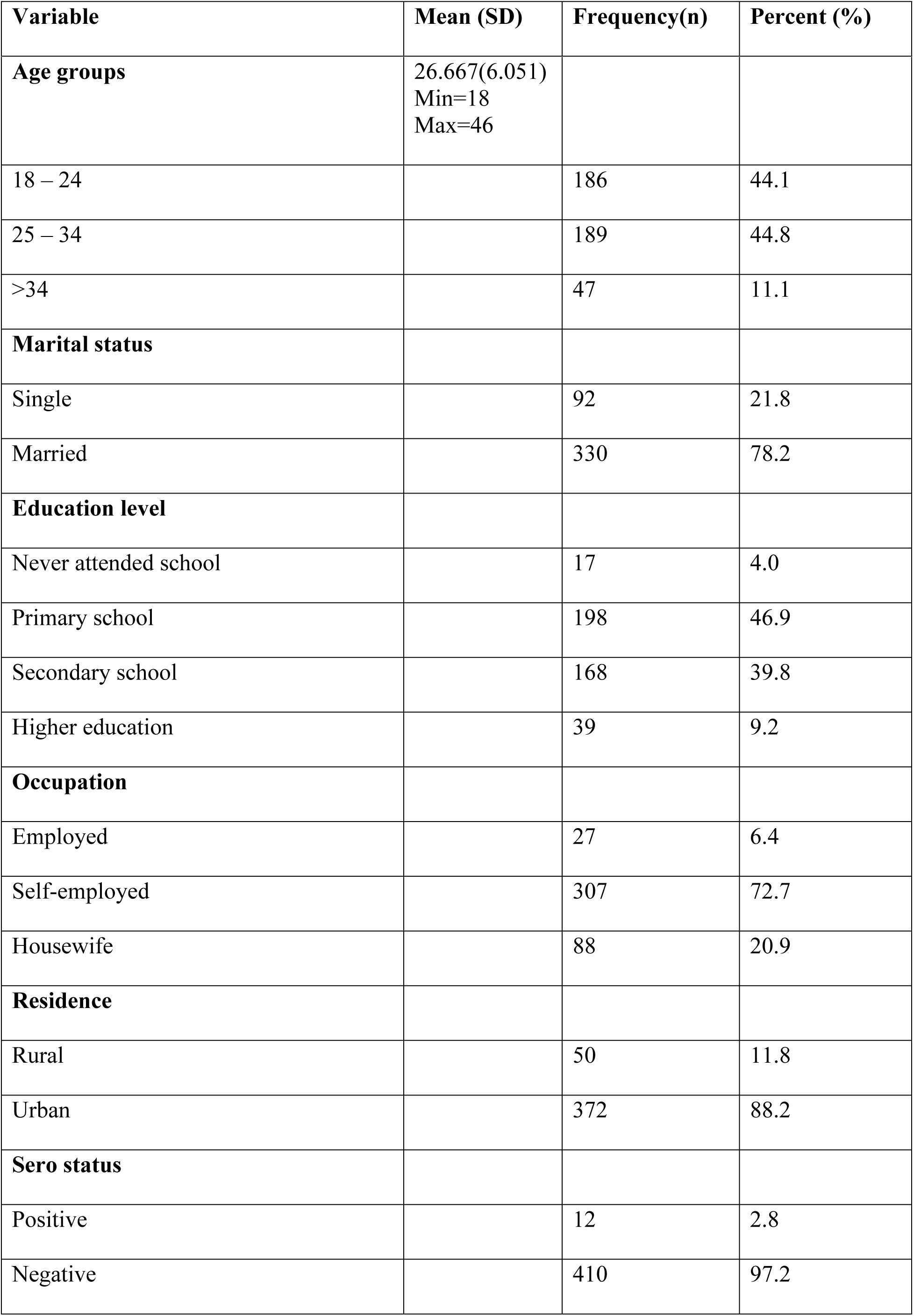
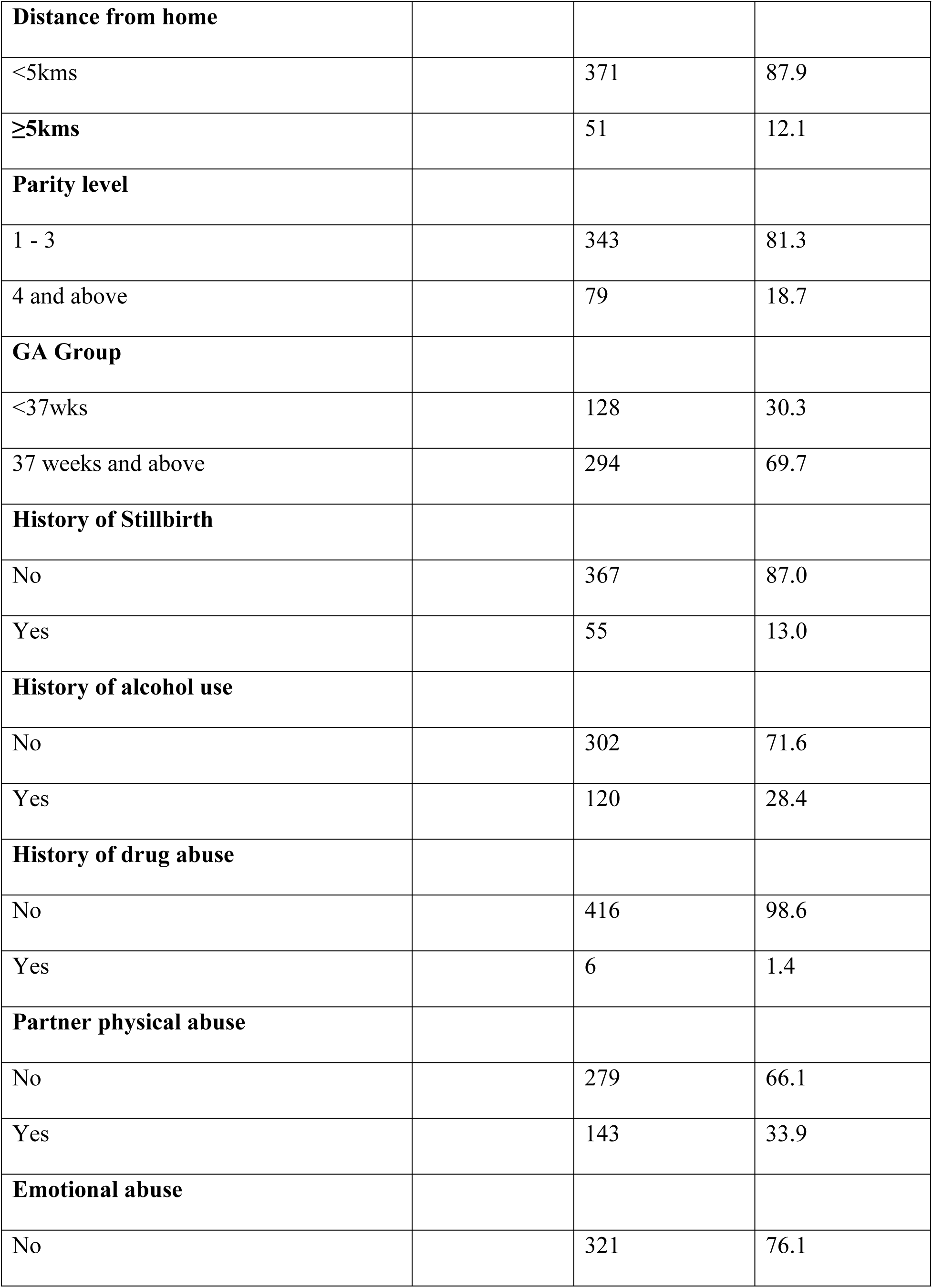
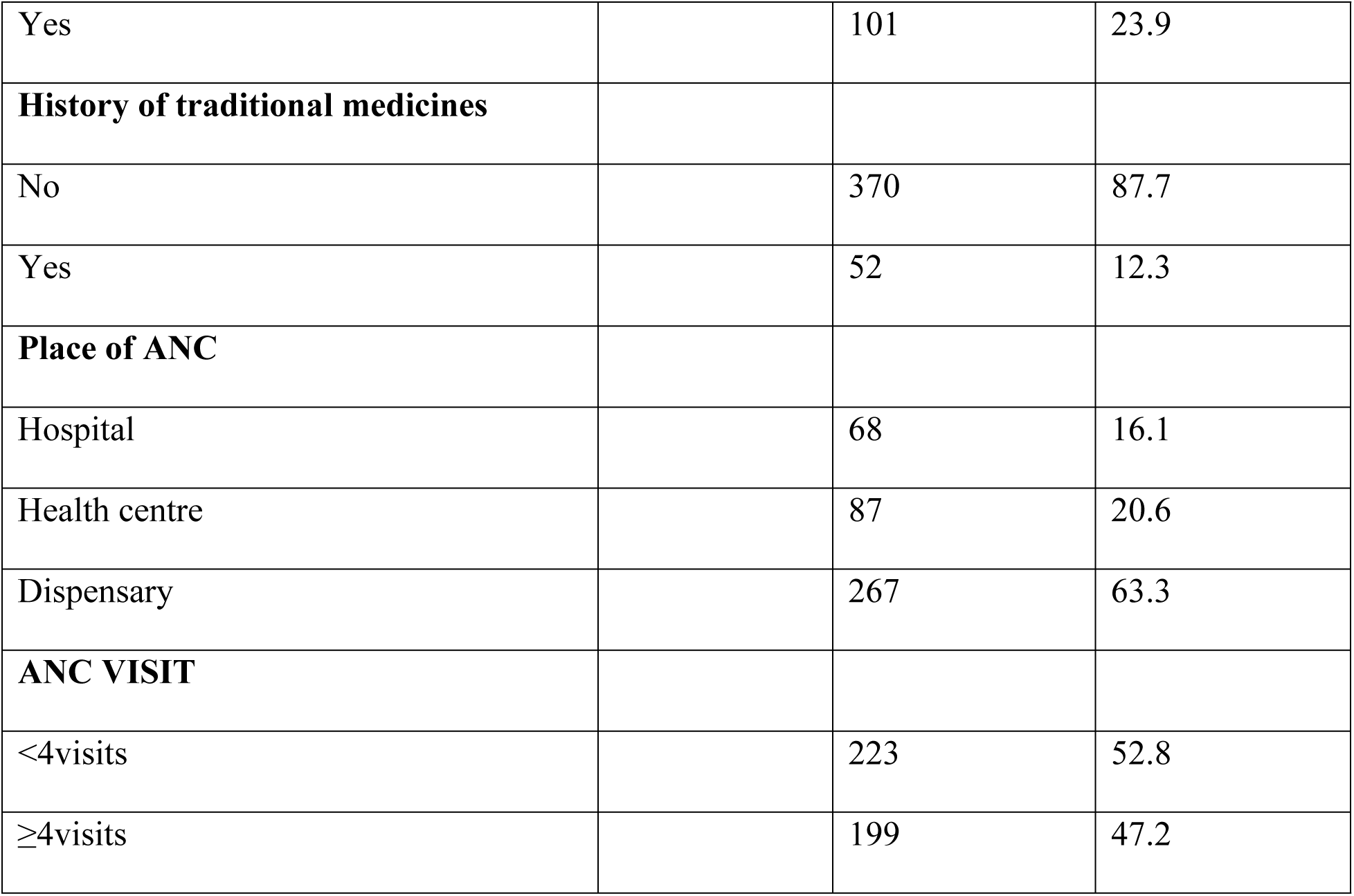
Background characteristics of participants (N = 422)

Within each facility, a systematic random sampling method was used to select study participants. The kth interval was obtained using a formula k^th^=N/n

Where kth =

N=Total estimated number of attendances per day

n =Total participants required per day

### Data Collection Methods and Tool

Two methods of data collection were used. The interviewer-administered questionnaire was used to obtain data on the background characteristics of postnatal mothers, and a documentary review using antenatal card number one and mother case note was used to collect information about obstetric information, neonatal information, and birth outcomes.

Data from the respondents in Dodoma selected health facilities were collected for two months, from April to May 2024, by the principal investigator and three diploma nurses as research assistants. Respondents were asked questions, and their responses were filled out in a questionnaire.

### Data Collection Tool

The paper based structured questionnaire with closed-ended questions adapted from the study conducted in Ethiopia was used for assessing sociodemographic, obstetric, maternal, and neonatal information, and adverse birth outcomes (21), and for assessing ANC services utilization was adapted from (22). The tool consists of five sections. Section A (Maternal sociodemographic characteristics), section B (Obstetric characteristics), section C (ANC service utilization), section D (Maternal adverse outcomes), and section E (Neonatal adverse outcomes). English language was used in collecting data from respondents and it took about 10 minutes to complete the questionnaire.

### Data Collection Procedure

The researcher used three licensed diploma nurses as research assistants. After completing two days of orientation on the tool contents and data collection procedure, the principal investigator collected the data and closely supervised the data collection process.

### Definition of variables

#### Dependent variable

Maternal adverse birth outcomes are defined as when the mother presents with postpartum hemorrhage, eclampsia, anemia, infection, maternal morbidity, and mortality.

#### Independent variable

Maternal sociodemographic characteristics (age, education, occupation, marital status, and residence), obstetric characteristics (parity, gestation age, history of stillbirth place where ANC service received), antenatal care services (number of ANC visits, timing, received ANC services), neonatal factors (age,body weight,apgar score,gestational age at delivery). Moderator variables (biological factors, psychological factors, lifestyle and behavioral factors, and geographical factors).

### Measurement of variables

#### Dependent variable

In this study, two outcomes which are maternal outcomes

#### Maternal adverse outcomes

The binary scale measured this variable using a single item, whether a woman is having any adverse birth outcome or not. If the woman had one or more adverse outcomes, it was coded “1,” while those mothers with no adverse outcomes were coded “0.”

#### Independent Variable

The independent variables were divided into maternal social demographic and obstetric characteristics and antenatal service utilization.

### Maternal sociodemographic and obstetrics characteristics

Maternal sociodemographic and obstetrics characteristics were measured by 14 items on different scales whereby age (numerical scale), marital status, occupation, place of residence, parity, gravidity, maternal serostatus, previous history of stillbirth, place of delivery, history of substance use, history of physical abuse, history of emotional abuse (nominal scale), and educational level of the mother (ordinal scale).

### Antenatal services received (documentary review)

In antenatal care services utilization, a total of 15 items were measured using a dichotomous scale. The tool was adapted from the previous study (21). The response used a binary scale to measure the response to the ANC service received, with “yes” indicating the mother received the service and “no” indicating the mother did not receive the service. There were a total of 38 points on the tool. An ANC service utilization score of 0–18 (0–49%) was considered inadequate ANC service utilization, a score of 19–30 (50–79%) was considered intermediate service utilization, and a score of 31–38 (≥80%) was considered adequate service utilization.

### Data Analysis

Data was processed by using IBM SPSS version 25, data were entered into the software, and a database was created, which was followed by data cleaning by using statistical checking for missing data. Data were tested for normality to determine the normal distribution of data by running frequencies. The model of analysis was descriptive of social demographic characteristics and prevalence by descriptive analysis and presented by using frequency and percentage. Chi-square was used to measure the relationship between independent and dependent variables. The P-value <0.05 was considered significant. Inferential analysis that includes binary logistic regression, multivariable logistic regression, and 95% CI was used to measure the strength of the association.

### Ethical considerations

Ethical clearance with reference number MA.No.MA.84/261/70/24 was obtained from the Research and Ethical Committee Board of the University of Dodoma. Permission to conduct a study in the respective health facility was obtained from the Dodoma region’s regional administrative secretary (RAS). Both verbal and written consent were sought from study participants after explaining the study objectives, benefits, and risks of their participation. Participants were free to withdraw from the study at any time they wished. To ensure confidentiality, the respondent’s information was kept in a locked cupboard and only accessible by the researcher and the University of Dodoma for academic purposes.

## Results

### Social demographic and obstetric characteristics of the participants

Table 1 presents the background characteristics of the respondents in this section. The mean age of participants was 26.67±6.051, the maximum age was 46 years, and the minimum age was 18 years. Of the 422 participants, the majority, 186 (44.1%), belonged to the age group of 24-35 years. At 330 (78.2%), married people outnumbered single people. Furthermore, the majority, 307 (72.7%), were self-employed. In terms of education, 198 individuals (46.9%) had completed their primary education, and the majority resided within a 5-kilometer radius of 371 individuals (87.9%). Of the 422 participants, the majority were attending ANC at nearby dispensaries, with 371 (87.9%) being seronegative. Moreover, on obstetric characteristics, the majority were giving birth at a gestation age of 37 weeks and above, the majority of postnatal women 367 (87%) had no history of stillbirth, 307 (71.6%) had no history of alcohol use, 416 (98.6%) had no history of drug abuse, 279 (66.1) had no partner physical abuse, mothers with history emotional abuse 321(76.1%), no history of traditional medicine 370 (87.7%), a place of attended ANC most of them they were attended dispensary.

### Services received during ANC

Table 2 reveals that 225 (53.3%) of the participants had fewer than four ANC visits, while 331 (78.4%) booked into an ANC clinic after 12 weeks gestation. Based on the services received during ANC visits, the results indicated that 361 (85.5%) of the participants received intermediate ANC services, while 34 (8.1%) received inadequate ANC services.

**Table 2:**
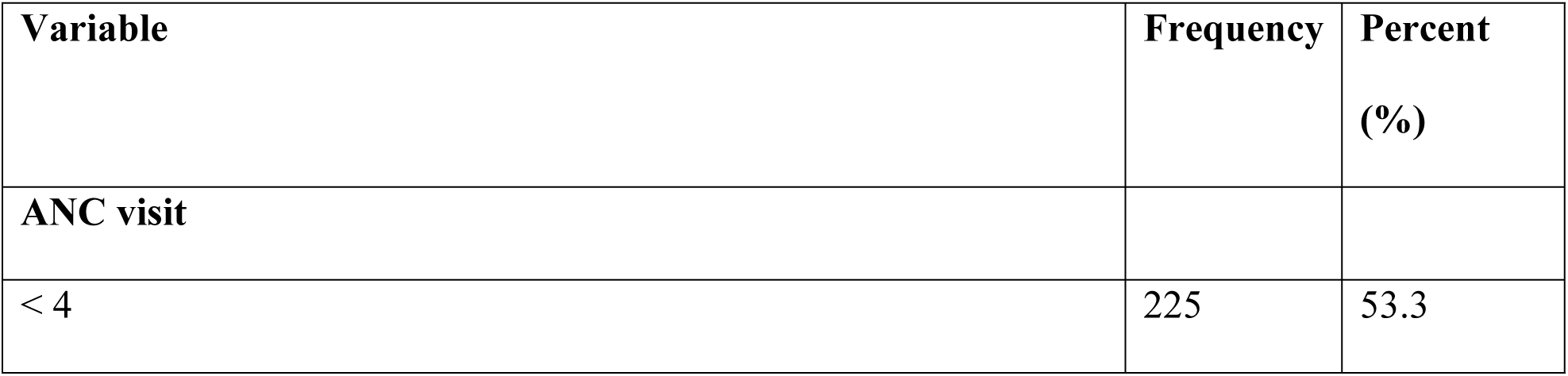
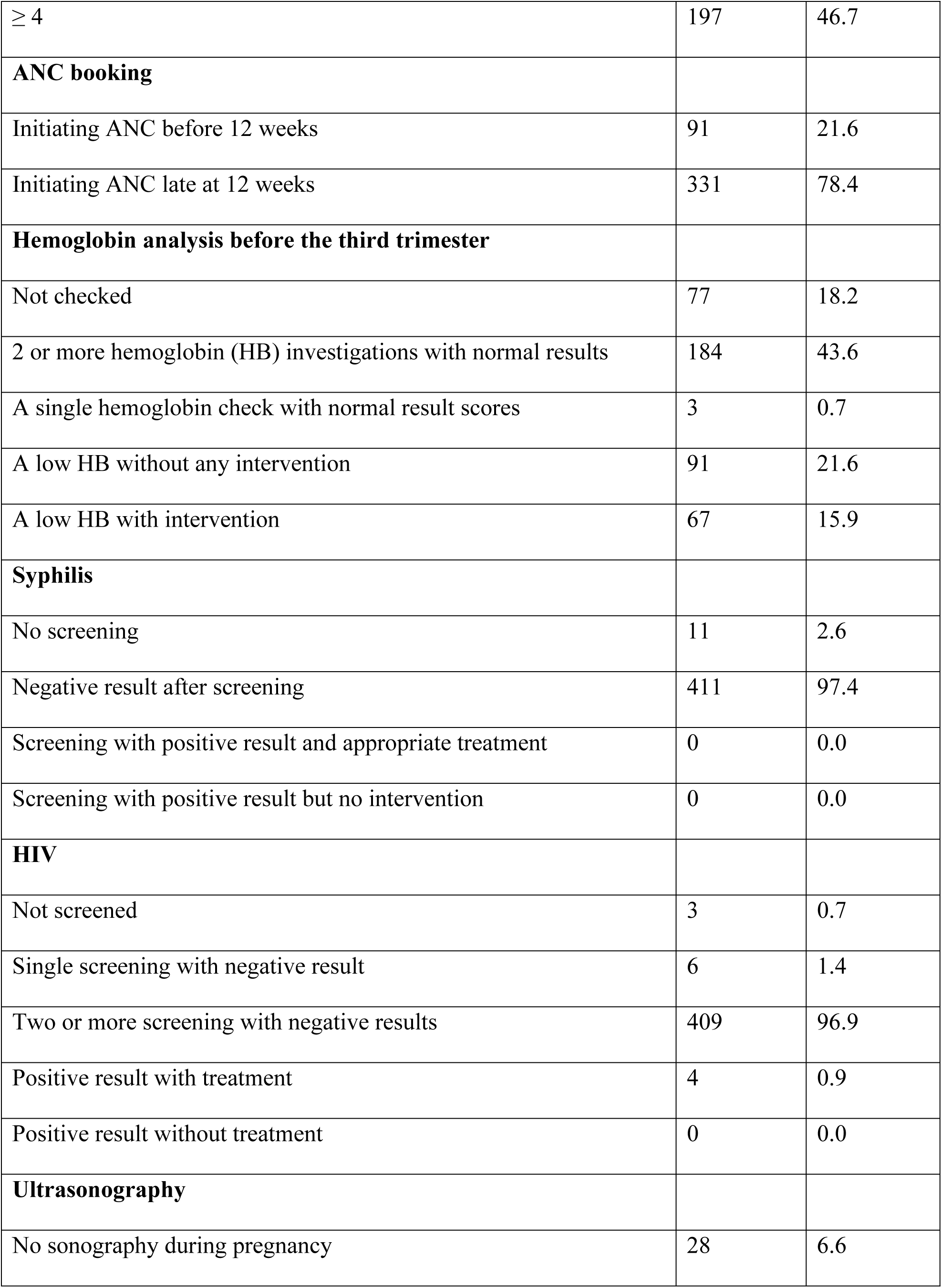
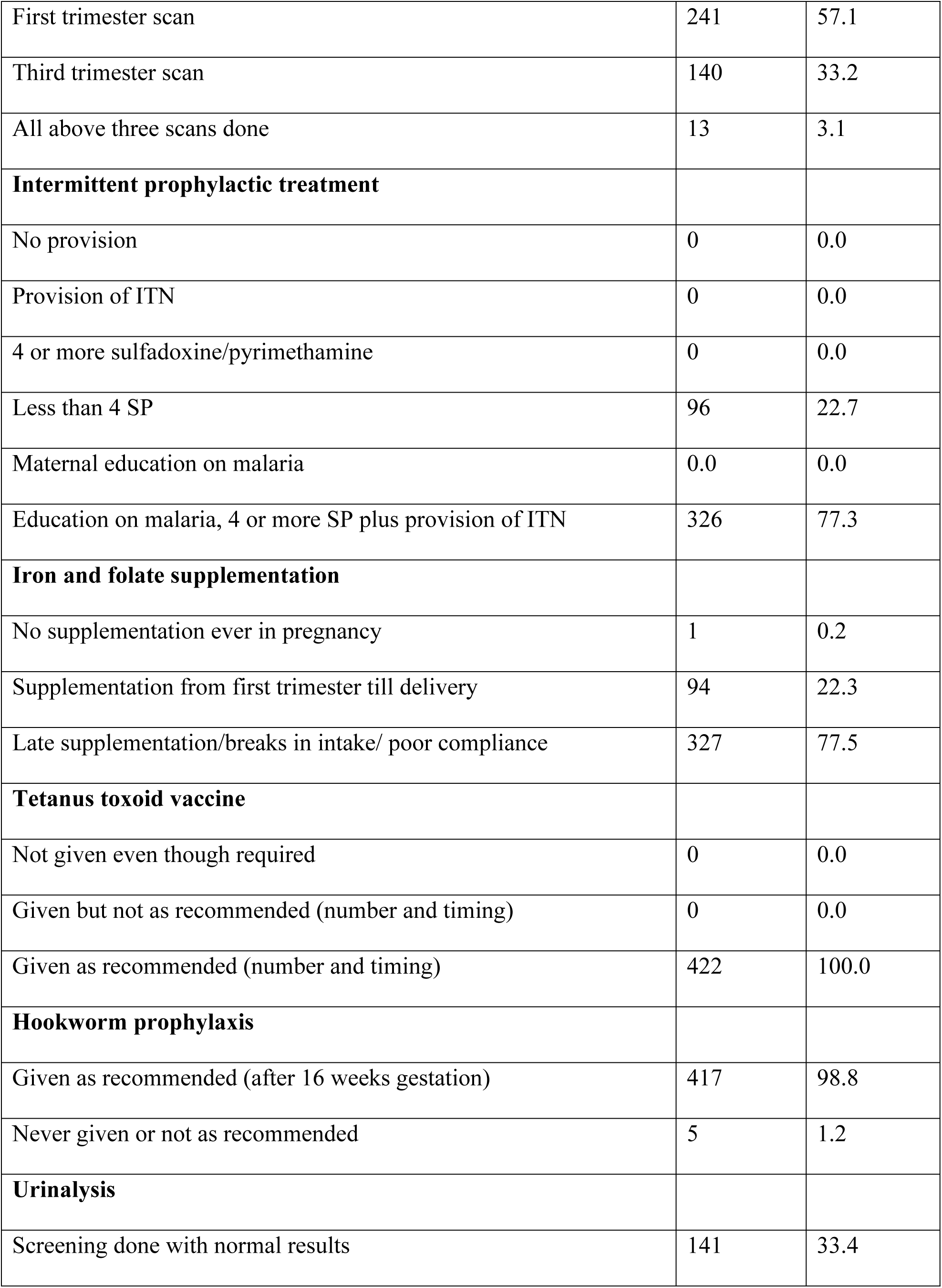
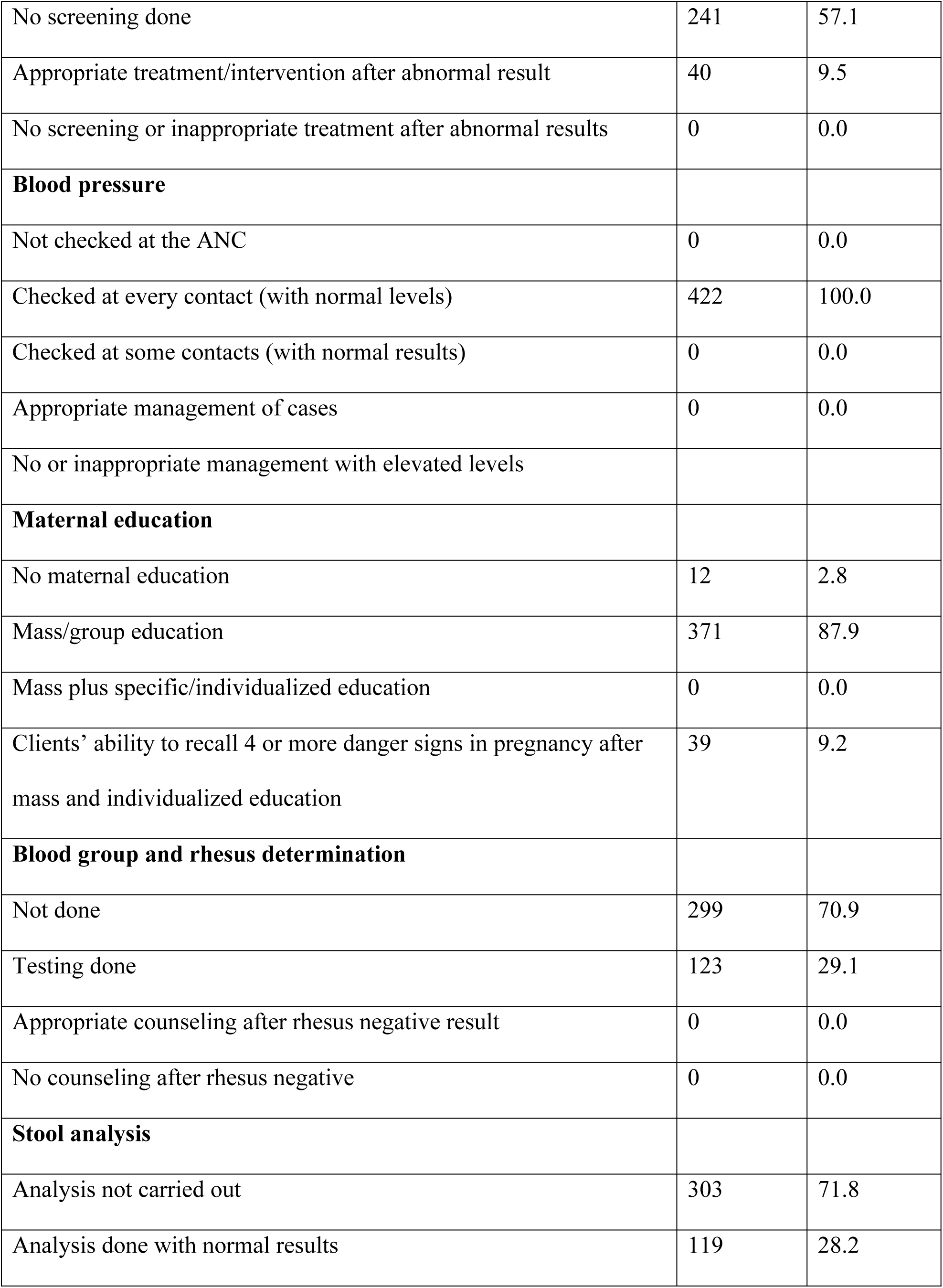
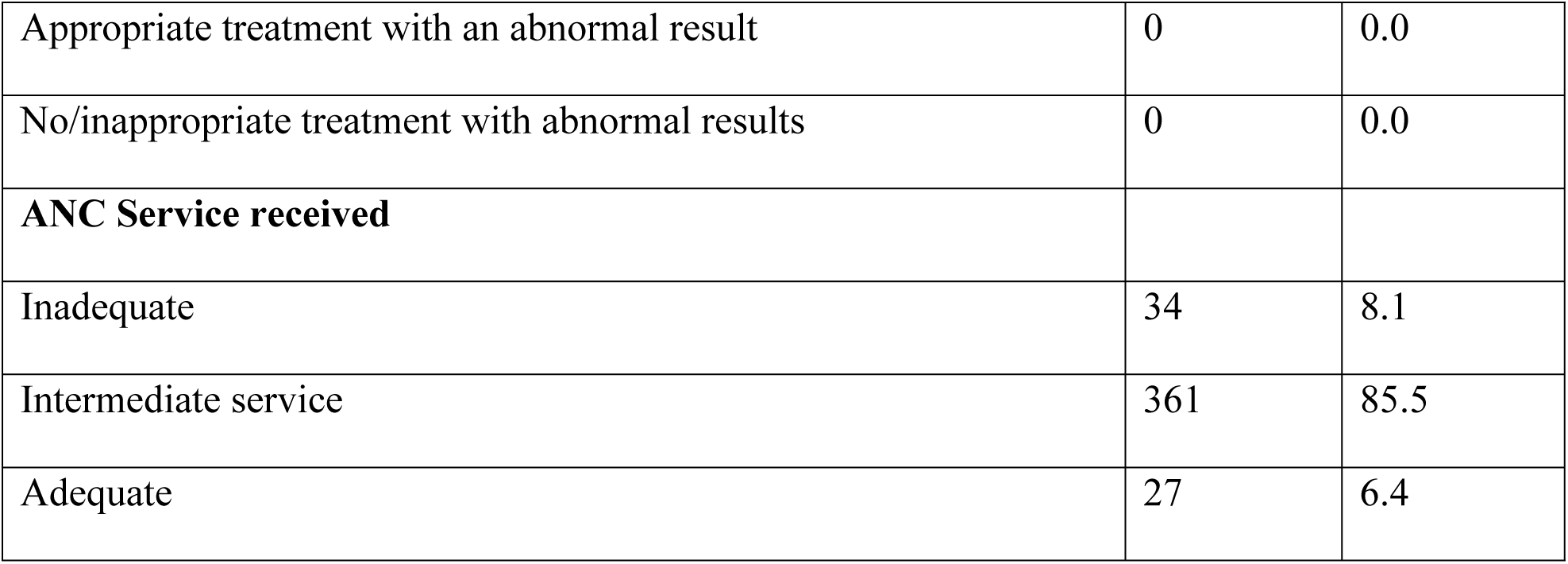
Item analysis on service received during ANC visits (N = 422)

| **Variable** | **Frequency** | **Percent**<br>**(%)** |
| --- | --- | --- |
| **ANC visit** |  |  |
| < 4 | 225 | 53.3 |
| ≥ 4 | 197 | 46.7 |
| **ANC booking** |  |  |
| Initiating ANC before 12 weeks | 91 | 21.6 |
| Initiating ANC late at 12 weeks | 331 | 78.4 |
| **Hemoglobin analysis before the third trimester** |  |  |
| Not checked | 77 | 18.2 |
| 2 or more hemoglobin (HB) investigations with normal results | 184 | 43.6 |
| A single hemoglobin check with normal result scores | 3 | 0.7 |
| A low HB without any intervention | 91 | 21.6 |
| A low HB with intervention | 67 | 15.9 |
| **Syphilis** |  |  |
| No screening | 11 | 2.6 |
| Negative result after screening | 411 | 97.4 |
| Screening with positive result and appropriate treatment | 0 | 0.0 |
| Screening with positive result but no intervention | 0 | 0.0 |
| **HIV** |  |  |
| Not screened | 3 | 0.7 |
| Single screening with negative result | 6 | 1.4 |
| Two or more screening with negative results | 409 | 96.9 |
| Positive result with treatment | 4 | 0.9 |
| Positive result without treatment | 0 | 0.0 |
| **Ultrasonography** |  |  |
| No sonography during pregnancy | 28 | 6.6 |
| First trimester scan | 241 | 57.1 |
| Third trimester scan | 140 | 33.2 |
| All above three scans done | 13 | 3.1 |
| **Intermittent prophylactic treatment** |  |  |
| No provision | 0 | 0.0 |
| Provision of ITN | 0 | 0.0 |
| 4 or more sulfadoxine/pyrimethamine | 0 | 0.0 |
| Less than 4 SP | 96 | 22.7 |
| Maternal education on malaria | 0.0 | 0.0 |
| Education on malaria, 4 or more SP plus provision of ITN | 326 | 77.3 |
| **Iron and folate supplementation** |  |  |
| No supplementation ever in pregnancy | 1 | 0.2 |
| Supplementation from first trimester till delivery | 94 | 22.3 |
| Late supplementation/breaks in intake/ poor compliance | 327 | 77.5 |
| **Tetanus toxoid vaccine** |  |  |
| Not given even though required | 0 | 0.0 |
| Given but not as recommended (number and timing) | 0 | 0.0 |
| Given as recommended (number and timing) | 422 | 100.0 |
| **Hookworm prophylaxis** |  |  |
| Given as recommended (after 16 weeks gestation) | 417 | 98.8 |
| Never given or not as recommended | 5 | 1.2 |
| **Urinalysis** |  |  |
| Screening done with normal results | 141 | 33.4 |
| No screening done | 241 | 57.1 |
| Appropriate treatment/intervention after abnormal result | 40 | 9.5 |
| No screening or inappropriate treatment after abnormal results | 0 | 0.0 |
| **Blood pressure** |  |  |
| Not checked at the ANC | 0 | 0.0 |
| Checked at every contact (with normal levels) | 422 | 100.0 |
| Checked at some contacts (with normal results) | 0 | 0.0 |
| Appropriate management of cases | 0 | 0.0 |
| No or inappropriate management with elevated levels |  |  |
| **Maternal education** |  |  |
| No maternal education | 12 | 2.8 |
| Mass/group education | 371 | 87.9 |
| Mass plus specific/individualized education | 0 | 0.0 |
| Clients' ability to recall 4 or more danger signs in pregnancy after mass and individualized education | 39 | 9.2 |
| **Blood group and rhesus determination** |  |  |
| Not done | 299 | 70.9 |
| Testing done | 123 | 29.1 |
| Appropriate counseling after rhesus negative result | 0 | 0.0 |
| No counseling after rhesus negative | 0 | 0.0 |
| **Stool analysis** |  |  |
| Analysis not carried out | 303 | 71.8 |
| Analysis done with normal results | 119 | 28.2 |
| Appropriate treatment with an abnormal result | 0 | 0.0 |
| No/inappropriate treatment with abnormal results | 0 | 0.0 |
| **ANC Service received** |  |  |
| Inadequate | 34 | 8.1 |
| Intermediate service | 361 | 85.5 |
| Adequate | 27 | 6.4 |

### Prevalence of maternal adverse outcomes (N=422)

The prevalence of maternal adverse outcome among post-natal mothers was 115(27.3%) (Figure 1)

**Figure 1.** Prevalence of maternal adverse outcomes

### Characterization of maternal adverse outcomes (N=115)

The distribution of maternal adverse outcomes was; hypertensive disorders 33, anemia 29%, postpartum hemorrhage 25%, and infections 13% (Figure 2.)

**Figure 2.** Shows the distribution of maternal adverse outcomes

### Bivariate and multivariate binary logistic analysis on factors associated with maternal adverse outcome

Bivariate and multivariate binary logistic regression to identify the factors solely influencing maternal adverse outcomes. The findings showed that participants who were emotionally abused were 1.766 times more likely to develop PPH, by 3.079 times more likely to develop eclampsia, and 4.275 times more likely to develop anemia compared to participants who were not emotionally abused (AOR = 3.864; 95%CI: 1.766, 8.456: *p* = 0.001), (AOR = 3.079; 95%CI: 1.514, 6.262: *p =* 0.002), and (AOR = 4.275; 95%CI: 2.020, 9.045: *p =* <0.001) respectively. Participants who had a history of stillbirth were 3.811 times more likely to develop eclampsia compared to participants who had no history of stillbirth (AOR = 3.811; 95%CI: 1.696, 8.562: *p = 0.001).* Participants who had their delivery conducted at GA <37 weeks were 2.846 times more likely to have anemia compared to participants who delivered with a GA ≥37 weeks (AOR = 2.846; 95%CI: 1.327, 6.102: *p* = 0.007). In addition, participants who had <4 ANC visits were 5.091 more likely to have an infection than those who had ≥4 ANC visits (AOR = 5.091; 95%CI: 1.093, 23.717: *p = 0.038)*

**Table 3:** The bivariate and multivariable binary logistic regression of factors associated with PPH, Eclampsia, Anaemia and Infection (N=422)

| Variable | Univariate |  |  |  | Multivariate |  |  |  |
| --- | --- | --- | --- | --- | --- | --- | --- | --- |
|  | COR | 95% CI |  | p-value | AOR | 95% CI |  | P-value |
|  |  | Lower | Upper |  |  | Lower | Upper |  |
| PPH |  |  |  |  |  |  |  |  |
| Residence |  |  |  |  |  |  |  |  |
| Rural | 2.243 | 0.799 | 5.359 | 0.134 | 2.327 | 0.860 | 6.297 | 0.096 |
| Urban | Ref |  |  |  | Ref |  |  |  |
| GA Group |  |  |  |  |  |  |  |  |
| <37wks | 1.964 | 0.915 | 4.214 | 0.83 | 1.756 | 0.800 | 3.855 | 0.161 |
| 37 weeks and above | Ref |  |  |  | Ref |  |  |  |
| Partner physical abuse |  |  |  |  |  |  |  |  |
| No | Ref |  |  |  | Ref |  |  |  |
| Yes | 1.910 | 0.895 | 4.077 | 0.094 | 1.812 | 0.831 | 3.952 | 0.135 |
| Emotional abuse |  |  |  |  |  |  |  |  |
| No | Ref |  |  |  | Ref |  |  |  |
| Yes | 3.825 | 1.777 | 8.232 | 0.001 | 3.864 | 1.766 | 8.456 | 0.001 |
| ECLAMPSIA |  |  |  |  |  |  |  |  |
| GA Group |  |  |  |  |  |  |  |  |
| <37wks | 1.766 | 0.894 | 3.488 | 0.101 | 1.620 | 0.791 | 3.320 | 0.187 |
| 37 weeks and above | Ref |  |  |  | Ref |  |  |  |
| History of alcohol use |  |  |  |  |  |  |  |  |
| No | 1.847 | 0.790 | 4.316 | 0.157 | 2.425 | 0.933 | 6.305 | 0.069 |
| Yes | Ref |  |  |  | Ref |  |  |  |
| **Emotional abuse** |  |  |  |  |  |  |  |  |
| No | Ref |  |  |  | Ref |  |  |  |
| Yes | 3.683 | 1.864 | 7.278 | <0.001 | 3.079 | 1.514 | 6.262 | **0.002** |
| **History of Stillbirth** |  |  |  |  |  |  |  |  |
| No | Ref |  |  |  | Ref |  |  |  |
| Yes | 2.115 | 0.911 | 4.908 | 0.081 | 3.811 | 1.696 | 8.562 | **0.001** |
| **History of traditional medicines** |  |  |  |  |  |  |  |  |
| No | Ref |  |  |  | Ref |  |  |  |
| Yes | 1.701 | 0.708 | 4.089 | 0.235 | 2.499 | 0.894 | 6.985 | 0.081 |
| **ANAEMIA** |  |  |  |  |  |  |  |  |
| **Age groups** |  |  |  |  |  |  |  |  |
| 18 – 24 | Ref |  |  |  | Ref |  |  |  |
| 25 – 34 | 1.853 | 0.831 | 4.128 | 0.131 | 2.039 | 0.868 | 4.790 | 0.102 |
| >34 | 2.095 | 0.680 | 6.454 | 0.198 | 2.908 | 0.878 | 9.635 | 0.081 |
| **GA Group** |  |  |  |  |  |  |  |  |
| <37wks | 2.661 | 1.299 | 5.452 | 0.007 | 2.846 | 1.327 | 6.102 | **0.007** |
| 37 weeks and above | Ref |  |  |  | Ref |  |  |  |
| **History of alcohol use** |  |  |  |  |  |  |  |  |
| No | Ref |  |  |  | Ref |  |  |  |
| Yes | 1.713 | 0.823 | 3.565 | 0.15 | 1.100 | 0.391 | 3.096 | 0.856 |
| **History of drug abuse** |  |  |  |  |  |  |  |  |
| No | Ref |  |  |  | Ref |  |  |  |
| Yes | 4.955 | 0.923 | 26.590 | 0.062 | 2.647 | 0.420 | 16.696 | 0.300 |
| **Partner physical abuse** |  |  |  |  |  |  |  |  |
| No | Ref |  |  |  | Ref |  |  |  |
| Yes | 1.942 | 0.950 | 3.968 | 0.069 | 1.751 | 0.653 | 4.699 | 0.266 |
| **Emotional abuse** |  |  |  |  |  |  |  |  |
| No | Ref |  |  |  | Ref |  |  |  |
| Yes | 4.424 | 2.139 | 9.152 | <0.001 | 4.275 | 2.020 | 9.045 | **<0.001** |
| **INFECTION** |  |  |  |  |  |  |  |  |
| **Occupation** |  |  |  |  |  |  |  |  |
| Employed | 6.960 | 0.606 | 79.957 | 0.119 | 4.622 | 0.339 | 63.005 | 0.251 |
| Self-employed | 3.539 | 0.454 | 27.600 | 0.228 | 2.603 | 0.322 | 21.012 | 0.369 |
| Housewife | Ref |  |  |  | Ref |  |  |  |
| **Residence** |  |  |  |  |  |  |  |  |
| Rural | 4.022 | 1.316 | 12.295 | 0.015 | 3.425 | 0.963 | 12.185 | 0.057 |
| Urban | Ref |  |  |  | Ref |  |  |  |
| **Parity level** |  |  |  |  |  |  |  |  |
| 1 - 3 | Ref |  |  |  | Ref |  |  |  |
| 4 and above | 3.050 | 1.053 | 8.836 | 0.040 | 3.155 | 0.964 | 10.320 | 0.057 |
| **History of alcohol use** |  |  |  |  |  |  |  |  |
| No | Ref |  |  |  | Ref |  |  |  |
| Yes | 2.277 | 0.807 | 6.424 | 0.120 | 0.959 | 0.234 | 3.930 | 0.953 |
| **Partner physical abuse** |  |  |  |  |  |  |  |  |
| No | Ref |  |  |  | Ref |  |  |  |
| Yes | 3.056 | 1.066 | 8.763 | 0.038 | 2.619 | 0.604 | 11.350 | 0.198 |
| **Emotional abuse** |  |  |  |  |  |  |  |  |
| No | Ref |  |  |  | Ref |  |  |  |
| Yes | 2.914 | 1.030 | 8.245 | 0.044 | 2.759 | 0.889 | 8.568 | 0.079 |
| **History of traditional medicines** |  |  |  |  |  |  |  |  |
| No | Ref |  |  |  | Ref |  |  |  |
| Yes | 2.720 | 0.833 | 8.881 | 0.097 | 1.645 | 0.380 | 7.118 | 0.505 |
| **ANC VISIT** |  |  |  |  |  |  |  |  |
| <4visits | 6.098 | 1.359 | 27.364 | 0.018 | 5.091 | 1.093 | 23.717 | **0.038** |
| ≥4visits | Ref |  |  |  | Ref |  |  |  |

## Discussion

This chapter presents a discussion of the study’s current findings, which aimed at assessing the prevalence and factors associated with adverse birth outcomes among postnatal mothers.

### Prevalence of maternal adverse outcomes

The findings of this study showed that the prevalence of maternal adverse outcomes was 27.3%. From 422 participants, postpartum hemorrhage 29 (25.2%), hypertensive disorders 38 (33.1%), postnatal anemia 33 (28.7%), and postnatal infection 15 (13%). The causes of this prevalence might be due to inadequate ANC check-ups up to less than four visits which limit some of the investigations, checks, and assessments. Moreover, postnatal mothers who didn’t receive adequate health education and failed to recall at least four sessions of health education were significantly associated with adverse maternal outcomes. However, this study is consistent with a study conducted in Uganda which reported a prevalence of maternal adverse outcomes of 37.6% (23).

This study supports that more than a quarter of respondents had less than four antenatal visits. Based on Tanzania ANC guideline recommendation every woman should be booking the first ANC services below twelve weeks of pregnancy and the recommended contact is eight contacts (24). Late ANC booking can delay diagnosis and receiving appropriate treatment and also will delay intervention of supplements and preventive measures. Nevertheless, late booking will increase the likelihood of risk such as anemia in pregnant.

This finding was relatively similar to the study conducted in Ethiopia which observed that the prevalence was 22.2% (25). Different study conducted in United Kingdom shows the prevalence of maternal adverse was 55%, the contributing factors was mothers breast infection (13.6%), abdominal pain (0.8%), and endometriosis (0.8%) (6). The differences in the prevalence of adverse maternal outcomes could be due to services delivery during prenatal care, including frequency of ANC attendance and services delivery and geographical location.

### Predictors associated with maternal adverse outcomes

Predictors found to be associated with maternal adverse outcomes among postnatal women were identified. Here are the maternal adverse outcomes with their predictors PPH (emotional abuse), anemia (emotional abuse, gestational age < 37 weeks). The current study reveals that mothers who experienced emotional abuse were more likely to develop PPH, but also mothers with anemia were those who experienced emotional abuse and who gave birth at a gestation age of less than 37 weeks.

These findings are similar to the study conducted in Ethiopia on the assessment of adverse pregnancy outcomes and their association factors (1), another study in Malawi on assessing factors associated with maternal mortality reported PPH and anemia were associated with emotional abuse and giving birth below 37 weeks gestation age (26). Factors such as delay in seeking delivery services, receiving care from lower health facilities, and inadequate skilled healthcare personnel, screening and treatment of existing health conditions like hypertension and anemia in pregnancy can contribute to maternal adverse outcomes. This study is different from the study conducted in Germany which shows predictors of PPH were those mothers with high BMI and substance abuse uses, dengue, heart disease, and lung disease (27). This could be due to different geographical locations and a sedentary lifestyle.

Likewise, mothers who got postpartum infection and eclampsia are those who attended ANC less than 4 visits and those who had a history of stillbirth and emotional abuse. These findings are consistent with a cohort study conducted in Western Nepal that yielded a similar result in which women with a history of stillbirth have an increased risk of eclampsia and postnatal infection (28). Insufficient intervention during antenatal care, lack of in-depth assessment, inadequate utilization of ANC services, and lack of health education about danger signs can contribute to the occurrence of postnatal infections and eclampsia.

This study is similar to the study conducted in Tanzania on assessing prevalence and factors associated with postnatal infection (29). Moreover, a study conducted shows a postnatal mother with a history of stillbirth and inadequate ANC visits are predictors of postpartum infection and eclampsia (1). This study is different from to study conducted in Nigeria reported causes of postnatal infection were those mothers who were delivered by the caesarian section(30). This difference is due to the model of delivery and different populations.

Postnatal mothers who experienced partner physical abuse were significantly associated with PPH, eclampsia, and anemia compared with those who did not experience partner physical abuse. Partner abuse significantly impacts maternal health and contributes to maternal adverse birth outcomes by facing difficulties in obtaining adequate nutrition, leading to essential vitamins and minerals monitoring of pregnancy and treatment of abnormal conditions. Poor nutrition status is a known risk factor for anemia during pregnancy. Furthermore, chronic stress due to partner abuse leads to unhealthy choices or neglecting health needs which exacerbates the risk of anemia and chronic diseases during pregnancy and postnatal anemia.

Moreover, women who have emotional abuse may delay medical care services for fear of their relatives. This delay significantly increases the risk of secondary complications, Also abusive relationships can lead to neglected personal hygiene and care increasing the risk of infection during pregnancy and post-delivery where this neglect includes inadequate prenatal care and lack of attention to signs of infection.

Due to maternal adverse outcomes, an emotional abuse screen of pregnant women against violence is necessary during antenatal care services to prevent maternal adverse outcomes through appropriate management (24).

This finding aligns with a Tanzanian study that revealed a higher likelihood of adverse maternal outcomes for postnatal women who endured emotional abuse from their relatives compared to those who did not have emotional abuse (31). This finding is inconsistent with a study conducted in Ethiopia (32).

Women who are single and pregnant, as well as those who are unemployed and dependent, were more likely to have maternal adverse outcomes due to emotional abuse. This psychological stress leads to a high level of stress hormones such as cortisol, which can also affect immune function work make women more susceptible to infection.

Mothers who had stress associated with emotional abuse were led to physical symptoms including fatigue and weakened immune response impacted their health-seeking behavior. The women who experienced emotional abuse were likely to seek prenatal care less than four visits which may lead to addressing health issues such as anemia and infection and the untreated condition is significantly associated with adverse birth outcomes due to untreated conditions.

Postnatal women who had fewer than four ANC visits were significantly associated with anemia compared with those who attended more than four visits. Findings are a bit in line with a study conducted on assessing factors that contribute to postnatal anemia in Ethiopia (33). Possible factors could be inadequate ANC services delivered, attending antenatal less than four visits were statistically associated with anemia due to poor monitoring and supplements during ANC visits.

### Study limitations

This study could not be generalized since it was conducted only in the Dodoma region. Also, this design does not establish a causal relationship because the cross-sectional design can identify correlations between factors and outcomes but cannot prove the causation does not include under-18 mothers because of ethical considerations, a study needed to involve minors requires additional ethical parental consent permits. Nevertheless, it was limiting the ability to assess long-term adverse outcomes as the data was collected at a single point in time

## Conclusion

The study findings indicate that adverse neonatal outcomes remain high, with a prevalence of 21%, and maternal adverse outcomes, with a prevalence of 27.3%. These results show that adverse birth outcomes still exist in the Dodoma region. Important factors that contribute to maternal adverse outcomes are those postnatal women who attended fewer than four visits, mothers with a history of stillbirth, mothers who experienced partner physical abuse; those who experienced emotional abuse, and those who were living more than five kilometers from home and the ANC facility.

## Data Availability

Ethical considerations were done.

## Competing Interest

Authors have declared there was no conflict of interest.

## Acknowledgment

The author would like to extend acknowledgment to the University of Dodoma for its support on the accomplishment of the study.

## Author Contributions

**Conceptualization:** Kina Shafii Msuya

**Data curation:** Kina Shafii Msuya

**Investigation:** Kina Shafii Msuya

**Formal analysis:** Kina Shafii Msuya, Fabiola V. Moshi, Walter C. Millanzi

**Methodology:** Kina Shafii Msuya, Fabiola V. Moshi, Walter C. Millanzi

**Supervision:** Fabiola V. Moshi, Walter C. Millanzi

**Writing original manuscript draft:** Kina Shafii Msuya, Fabiola V. Moshi, Walter C. Millanzi

**Writing reviewing & editing:** Kina Shafii Msuya, Fabiola V. Moshi, Walter C. Millanzi

## Notes

### Competing Interest Statement

The authors have declared no competing interest.

### Funding Statement

No funding for submission

### Author Declarations

Ethical clearance with reference number MA.No.MA.84/261/70/24 was obtained from the Research and Ethical Committee Board of the University of Dodoma

